# COVID-19 vaccination rates among health care workers by immigrant background. A nation-wide registry study from Norway

**DOI:** 10.1101/2021.09.17.21263619

**Authors:** Kristian Bandlien Kraft, Ingeborg Elgersma, Trude Marie Lyngstad, Petter Elstrøm, Kjetil Telle

## Abstract

**Background:** Studies have suggested that some minority groups tend to have lower vaccination rates than the overall population. This study aims to examine COVID-19 vaccination rates among health care workers (HCWs) in Norway, according to immigrant background.

**Methods:** We used individual-level, nation-wide registry data from Norway to identify all HCWs employed full-time at 1 December 2020. We examined the relationship between country of birth and COVID-19 vaccination from December 2020 to August 2021, both crude and adjusted for e.g. age, sex, municipality of residence, and detailed occupation codes in logistic regression models.

**Results:** Among all HCWs in Norway, immigrants had a 9 percentage point lower vaccination rate (85%) than HCWs without an immigrant background (94%) at 31 August 2021. The overall vaccination rate varied by country of birth, with immigrants born in Russia (71%), Serbia (72%), Lithuania (72%), Romania (75%), Poland (76%), Eritrea (77%), and Somalia (78%) having the lowest crude vaccination rates. When we adjusted for demographics and detailed occupational codes, immigrant groups that more often worked as health care assistants, such as immigrants from Eritrea and Somalia, increased their vaccination rates.

**Conclusion:** Substantial differences in vaccination rates among immigrant groups employed in the health care sector in Norway indicate that measures to improve vaccine uptake should focus specific immigrant groups rather than all immigrants together. Lower vaccination rates in some immigrant groups appears to be largely driven by the occupational composition, suggesting that some of the differences in vaccine rates can be attributed to variation in vaccine access.

## Introduction

As of 5 September 2021, 91% of health care workers (HCWs) in Norway had received either one or two doses of a vaccine against SARS-CoV-2, the virus causing coronavirus disease 2019 (COVID-19).^1^ High vaccine uptake in a population is crucial for vaccine effectiveness. Furthermore, a high vaccine uptake among HCWs is vital to secure the operation of the health care services and the health of vulnerable patients.

Studies have suggested that some minority groups tend to be more vaccine hesitant than majority groups.^2-4^ A survey in Norway on attitudes toward COVID-19 vaccines indicated that individuals born in Eastern Europe, Western Asia and Africa tended to be more vaccine hesitant.^2^ A study from the United Kingdom (UK), using representative survey data collected between 24 November and 1 December 2020 showed that vaccine hesitancy was more prevalent among Black, Pakistani/Bangladeshi, and mixed non-UK/Irish White ethnic groups.^3^ A systematic review over observational studies about the pandemic influenza vaccine H1N1 in the US showed that African American and Latinos had disproportionally lower vaccination rates than other groups.^4^ Together, these surveys indicate that there may be larger vaccine hesitancy in certain immigrant groups, possibly resulting in a lower vaccine uptake.

Reports from the COVID-19 vaccination program in European countries have indicated that there are differences in vaccination rates based on country of origin.^5-7^ A Norwegian report from April 2021 showed that persons above 75 years born in Norway, Sweden and Denmark had the highest vaccination rates (≥90%), while persons born in Somalia had the lowest vaccination rates (34%).^5^ Similar, the Public Health Agency of Sweden reported in April 2021 that for people aged 80 years and older, vaccination rates for non-immigrants were at 91%, versus 59% and 44% for people born in North Africa and other African countries.^6^ A preprint study on HCWs in UK find that a larger proportion of white HCWs have received a vaccine against COVID-19 than HCW with another ethnicity.^7^

To improve knowledge on vaccination in minority groups, this study aims to examine registered COVID-19 vaccination among immigrant groups across HCWs in Norway from December 2020 to August 2021.

The study focuses on HCWs, since they have been prioritised in the COVID-19 vaccination programme and are vital in securing the operation of the health care services and the health of vulnerable patients. The Norwegian vaccination program has prioritised essential HCWs who are critically hard to replace, or HCW with direct patient contact.^8^ However, a study finds that hesitancy may exists in a great extent among HCWs in Turkey.^9^ At 31 August, all HCWs should have been offered the COVID-19 vaccine. Since there may be differences in demographic and socioeconomic composition for immigrant groups, which in turn may be related to differences in vaccine uptake, we also adjust for age, sex, and detailed occupational groups.

## Methods

We have utilised individual-level data from BeredtC19, which is an emergency preparedness register aimed to provide rapid knowledge during the COVID-19 pandemic.^10^ The register was established by The Norwegian Institute of Public Health (NIPH) in cooperation with the Norwegian Directorate of Health and contains individual-level data for all Norwegian residents. The data in BeredtC19 utilized in this study, originated from the Norwegian Immunisation Registry (SYSVAK), vaccine type and vaccination date; the Register of Employers and Employees (AA-registeret), employment status and occupation; the Norwegian Population Registry, demographic information (sex, birth year, municipality of residence); registries maintained by Statistics Norway (SSB), immigrant status and country of birth; and the Norwegian Surveillance System for Communicable Diseases (MSIS), positive polymerase chain reaction (PCR) tests and serology tests for antibodies for SARS-CoV-2 from all laboratories in Norway. This individual-level data was linked across sources and over time using an encrypted version of the unique personal identification number given to every Norwegian resident at birth or upon immigration.

The establishment of BeredtC19 forms part of the legally mandated responsibilities of NIPH during epidemics. Institutional board review was conducted, and The Ethics Committee of South-East Norway confirmed (June 4th 2020, #153204) that external ethical board review was not required

### Study population

The study population included all HCWs in Norway with a contract of equal or more than hours per week and aged between 20 and 65 at 1^st^ of December 2020. Persons who were not permanent residents of Norway longer than six months (tourists, asylum applicants, etc.) could not be included. Additionally, we excluded persons without known municipality of residence (n=8). We identified HCWs following Molvik et al. in using ISCO-08 4-digit occupation codes, in combination with standard industrial classifications from “AA-registeret” (see Molvik et al. for details, and supplementary Table 2a and b for the detailed codes).^11^ The study population consists therefore of workers with patient-contact, such as physicians, nurses and health care assistants, as well as cleaning staff, but not administrative or kitchen staff. In total, the study population numbers 356 053 persons.

### Definitions of population groups

The residents of Norway were divided into three groups based on definitions by Statistics Norway:

- *immigrants:* persons residing in Norway, but born outside of Norway with two parents born outside Norway
- *Norwegian-born to immigrant parents*: persons born in Norway with two parents born outside Norway
- *rest of the population*, also referred to as *persons without immigrant background*: persons not belonging to the groups *immigrants* or *Norwegian-born to immigrant parents*

In some of the analyses we disaggregate the immigrant group into groups based on country of birth. We have focused on immigrant groups with at least 1000 HCWs in our data *or* with at least 15 000 residents in Norway. This yields the following 26 countries: Poland, Lithuania, Somalia, Pakistan, Sweden, Syria, Iraq, Eritrea, Germany, The Philippines, Vietnam, Iran, Thailand, Russia, Afghanistan, Denmark, Turkey, India, Bosnia-Hercegovina, Romania, Kosovo, Great Britain, Sri Lanka, Ethiopia, United States of America (USA), and Serbia. When the results are disaggregated by country of birth, *Norwegian-born to immigrant parents* are not included in their parents’ country of birth, but with the rest of the population. When reporting results for immigrants as one group, we include all immigrants (i.e. not only those in the 26 countries).

### Outcome Variable: COVID-19 vaccination

The outcome is a binary variable capturing if the person had received at least one dose of a vaccine against SARS-CoV-2 in Norway from 28 December 2020 to 31 August 2021.

### Explanatory variables

Among HCWs, timing of access to COVID-19 vaccines differed across occupations, and HCWs are therefore divided into six occupational groups (physicians, specialist nurses, nurses, nursing associates, health care assistants and others). In the regression analyses we used more detailed categories (physicians, specialist nurses, nurses, nursing associates, health care assistants, ambulance personnel, cleaners, radiographers (and associated staff), bioengineers, social educators, dentists, occupational therapists, physiotherapists, psychologists, and other health care workers). The other included categorical variables were operationalized as follows: sex (male/female), age (<29, 30-39, 40-49, 50-59, >60), registered PCR-positive test (yes/no), COVID-19 infection (yes/no), medical risk group for severe COVID-19 (yes/no), institution of employment (hospital, other specialist health service, primary health care service (excl. nursing homes), home care service, nursing home, or other health service), and municipality of residence (356 municipalities of Norway).

### Statistical methods

Since the outcome variable is binary, we employed logistic regression models with robust standard errors. From the logistic model we calculated the predicted probabilities of receiving a COVID-19 vaccine. The following three models were estimated: (1) The crude model, simply the share in each group vaccinated; (2) the partially adjusted model, the share of each group vaccinated adjusted for age, sex, municipality of residence; and (3) the full model, the share of each group vaccinated adjusted for age, sex, municipality of residence, COVID-19 infection, medical risk group, occupation, and health care service of employment. For three of the municipalities there was no variation in the outcome variable (100% vaccination rate), resulting in automatically being dropped from the logistic regression model. Hence, the three municipalities were excluded from the whole sample (160 observations in total).

## Results

### Description of sample

We included 356 053 HCWs aged between 20 and 65 years, mean (SD) age 41 (13). 81% were women and 11% were living in the capital Oslo. Immigrant HCWs numbered 62 339 and had a similar mean age at 41 (11), a lower percent of females (76%) and a higher share living in Oslo (19%). There were 5 477 *Norwegian-born to immigrant parents* with a considerably lower mean age at 29 (8) and higher percentage living in Oslo (41%). Descriptive statistics on the sample including immigrants by country of birth are presented in the supplementary table 1.

### Crude vaccination rates

Table 1 presents the crude vaccination rates by immigrant background and occupation. Overall, 92% of all HCWs had been vaccinated by the end of our observation period on 31 August 2021 (Table 1). HCWs without immigrant background had the highest vaccination rate (94%). Immigrants and *Norwegian-born to immigrant parents* had an overall vaccination rate of 85% and 88%. This pattern holds across occupational groups, although the sizes of the differences vary with occupation.

**Table 1:** Frequency (N) and share (%) vaccinated with at least one dose of COVID-19 vaccine, by immigrant group and health care occupation on 31 August 2021.

| Immigrant background | All |  | Physicians |  | Specialist nurses |  | Nurses |  | Nursing associates |  | Health care assistants |  | Other health care workers |  |
| --- | --- | --- | --- | --- | --- | --- | --- | --- | --- | --- | --- | --- | --- | --- |
|  | N | % | N | % | N | % | N | % | N | % | N | % | N | % |
| All | 356 053 | 92 | 22 432 | 97 | 30 431 | 97 | 58 745 | 93 | 90 675 | 91 | 91 086 | 89 | 60 659 | 94 |
| Persons without immigrant background | 287 818 | 94 | 16 779 | 98 | 27 667 | 97 | 50 303 | 94 | 70 590 | 93 | 70 520 | 91 | 50 389 | 95 |
| Immigrants | 62 546 | 85 | 4 761 | 95 | 2 66 | 93 | 7 882 | 85 | 19 097 | 85 | 18 738 | 81 | 9 167 | 88 |
| Norwegian-born to immigrant parents | 5 529 | 88 | 892 | 96 | 104 | 97 | 560 | 88 | 988 | 84 | 1 828 | 86 | 1 103 | 89 |
| Norway | 293 347 | 94 | 17 671 | 98 | 27 771 | 97 | 50 863 | 94 | 71 578 | 92 | 72 348 | 91 | 51 492 | 95 |
| Denmark | 1 29 | 95 | 211 | 97 | 200 | 96 | 207 | 92 | 178 | 93 | 174 | 91 | 308 | 98 |
| Sweden | 2 99 | 94 | 357 | 98 | 488 | 97 | 476 | 95 | 661 | 91 | 499 | 87 | 489 | 94 |
| Poland | 3 644 | 76 | 271 | 94 | 187 | 81 | 649 | 78 | 631 | 75 | 1 12 | 69 | 781 | 77 |
| Romania | 733 | 75 | 89 | 88 | 22 | - | 160 | 73 | 143 | 74 | 195 | 72 | 121 | 72 |
| Lithuania | 1 795 | 72 | 125 | 92 | 58 | 86 | 528 | 76 | 241 | 76 | 580 | 66 | 249 | 65 |
| UK | 370 | 94 | 54 | 94 | 28 | - | 43 | - | 67 | 96 | 99 | 92 | 76 | 97 |
| Russia | 1 873 | 71 | 211 | 86 | 65 | 83 | 248 | 74 | 554 | 68 | 571 | 64 | 214 | 77 |
| Turkey | 473 | 88 | 25 | - | 13 | - | 37 | - | 121 | 87 | 171 | 88 | 105 | 88 |
| Germany | 2 464 | 88 | 676 | 97 | 187 | 94 | 365 | 85 | 327 | 79 | 464 | 80 | 426 | 90 |
| Bosnia-Herzegovina | 1 222 | 85 | 135 | 95 | 60 | 85 | 148 | 86 | 436 | 81 | 230 | 81 | 204 | 89 |
| Serbia | 1 453 | 72 | 164 | 85 | 17 | - | 120 | 78 | 877 | 69 | 182 | 65 | 92 | 85 |
| Kosovo | 975 | 85 | 28 | - | 23 | - | 97 | 85 | 371 | 87 | 293 | 82 | 161 | 88 |
| Eritrea | 3 956 | 77 | 14 | - | 10 | - | 126 | 87 | 1 572 | 80 | 1 931 | 72 | 303 | 85 |
| Ethiopia | 1 793 | 85 | 26 | - | 24 | - | 155 | 88 | 768 | 84 | 644 | 84 | 173 | 85 |
| Somalia | 2 884 | 78 | 31 | - | 33 | - | 256 | 75 | 991 | 81 | 1 212 | 74 | 355 | 82 |
| Afghanistan | 1 757 | 87 | 65 | 98 | 12 | - | 161 | 87 | 536 | 89 | 813 | 85 | 156 | 89 |
| Sri Lanka | 1 438 | 95 | 106 | 97 | 61 | 93 | 139 | 92 | 469 | 95 | 289 | 94 | 374 | 98 |
| Philippines | 6 004 | 93 | 18 | - | 108 | 94 | 933 | 93 | 2 972 | 93 | 1 347 | 93 | 624 | 94 |
| India | 973 | 94 | 104 | 97 | 49 | - | 158 | 96 | 406 | 94 | 143 | 92 | 111 | 89 |
| Iraq | 1 476 | 87 | 170 | 98 | 21 | - | 98 | 88 | 359 | 87 | 573 | 82 | 243 | 90 |
| Iran | 1 846 | 92 | 240 | 97 | 111 | 96 | 212 | 89 | 437 | 93 | 486 | 88 | 342 | 94 |
| Pakistan | 909 | 91 | 200 | 95 | 14 | - | 47 | - | 169 | 93 | 274 | 87 | 197 | 90 |
| Syria | 744 | 84 | 47 | - | <5 | - | 8 | - | 155 | 85 | 481 | 83 | 50 | 80 |
| Thailand | 2 354 | 96 | 9 | - | 23 | - | 108 | 95 | 724 | 95 | 874 | 95 | 616 | 98 |
| Vietnam | 803 | 97 | 69 | 99 | 39 | - | 113 | 96 | 189 | 97 | 156 | 94 | 226 | 98 |
| USA | 284 | 90 | 50 | - | 24 | - | 43 | - | 37 | - | 88 | 83 | 41 | 90 |
| Remaining immigrants | 16 043 | 85 | 1 266 | 94 | 781 | 93 | 2 247 | 83 | 4 706 | 85 | 4 849 | 82 | 2 13 | 88 |

Note: \*=Specialist nurses includes midwives. \*\*=Others includes ambulance personnel, cleaners, radiographers (and associated staff), bioengineers, social educators,
dentists, occupational therapists, physiotherapists, psychologists and other health care workers. The share vaccinated is hidden for cells with 50 persons and less due to anonymity considerations.

The lowest vaccination rates were among HCWs from Russia (71%), Serbia (72%), Lithuania (72%), Romania (75%), Poland (76%), Eritrea (77%), and Somalia (78%). The immigrant groups with the highest vaccination rate were from Vietnam (97%), Thailand (96%), Sri Lanka (95%), Denmark (95%), UK (94%), and India (94%).

In general, physicians (97%) and specialist nurses (97%) had the highest vaccination rate, while health care assistants (89%) had the lowest. The tendency for vaccination rates to be higher for higher-educated HCW, has been evident since the start of vaccination in Norway in January 2021, with the differences declining somewhat in the last month (see supplement figure 1). The pattern can also be observed for within several of the immigrant groups. Physicians and health care assistants from Lithuania, for example, have a vaccination rate of 92% and 66%, while physicians and health care assistants from Pakistan have a vaccination rate of 94% and 87%. How many HCWs who worked as health care assistants varied by country of birth (Figure 1). Several of the immigrant groups, especially those from Syria, Eritrea, Afghanistan and Somalia, had a disproportionally large share of the persons working as health care assistants. In the whole population 25% of HCWs were health care assistants, while the share of HCWs who work as health care assistants among immigrants from Syria, Eritrea, Afghanistan and Somalia were 65%, 49%, 46% and 42%.

**Figure 1:**
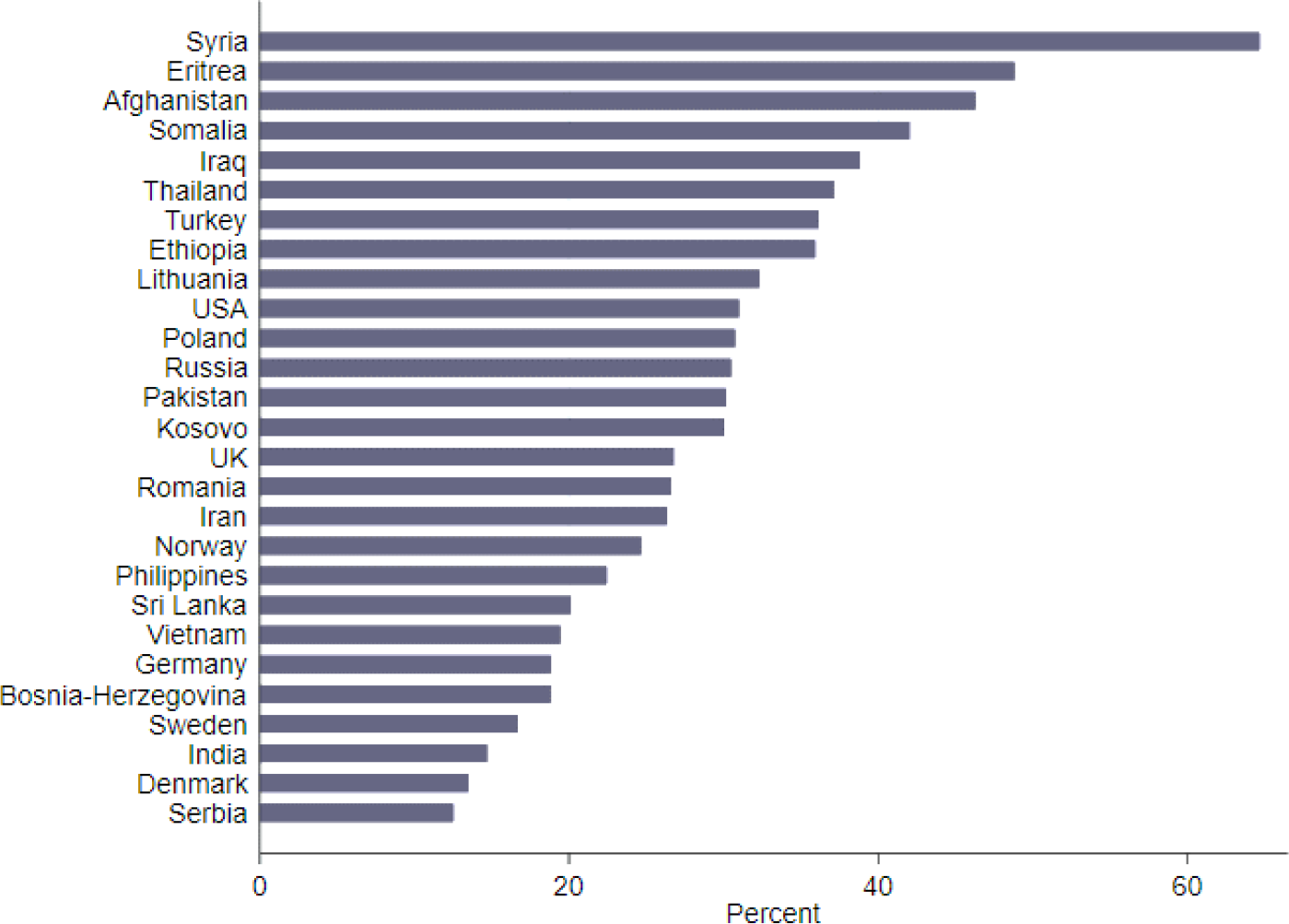
Share of HCWs that are employed as health care assistants by country of birth. In Norway on 1 December 2020.

### Adjusted vaccination rates

Both immigrants and *Norwegian-born to immigrant parents* had lower vaccination rates than *persons without immigrant background* in the crude model and in the adjusted models (Figure 2). Although the adjustments increase the likelihood that immigrants and *Norwegian-born to immigrant parents* are vaccinated, the differences are not completely attenuated. The full model predicts a larger probability for immigrants to be vaccinated than in the crude model – from 0.85 (95% confidence interval (CI) 0.851-0.856) to 0.88 (95% CI 0.877-0.882). This pattern can also be observed for *Norwegian-born to immigrant parents*, although it is more uncertain because of low numbers – from 0.88 (95% CI 0.874-0.891) to 0.90 (95% CI 0.895-0.910) (Figure 2). The adjustments in the partly adjusted model does not appear to affect the probability for immigrants, but in a larger extent for *Norwegian-born to immigrant parents*. It is likely that the young age composition of *Norwegian-born to immigrant parents* affect relative change in probability in the partly adjusted model.

**Figure 2:**
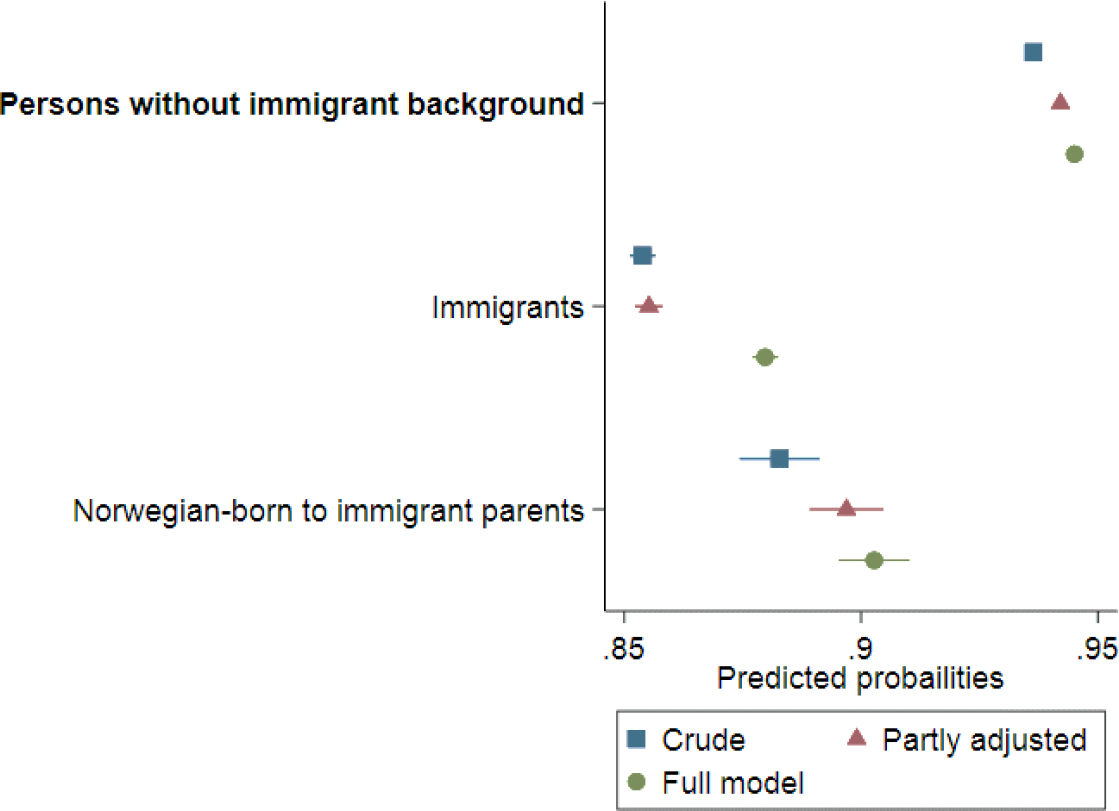
Predicted probability of COVID-19 vaccination by immigrant background on 31 August 2021, with 95% confidence intervals, based on results from three logistic regression models: Crude, partly and fully adjusted. Reference category: Persons without immigrant background (bold). Partly adjusted model: adjusted for age, sex and municipality. Full model: adjusted for age, sex, municipality, COVID-19, risk group for severe COVID-19, occupation and health care service of employment.

There were large variations between vaccination rates among HCWs based on country of birth (Figure 3). In general, the adjusted models predict a largely similar picture as the crude vaccination rates, with some exceptions. The immigrant groups with the lowest vaccination rates in the crude model have also the lowest predicted probability of vaccination in the adjusted models, and vice versa. However, when adjusting for all covariates in the full model the predicted probabilities of vaccination for immigrants from Eritrea and Somalia increase substantially: from 0.770 and 0.776 in the crude model, to 0.849 and 0.854 in the full model. The other immigrant groups with the lowest vaccination rates (Russia, Serbia, Lithuania, Romania and Poland) do not show a similarly large increase, although some do show a minor rise. All other immigrant groups show a similar pattern, with either a small increase in the predicted probability or no increase.

**Figure 3:**
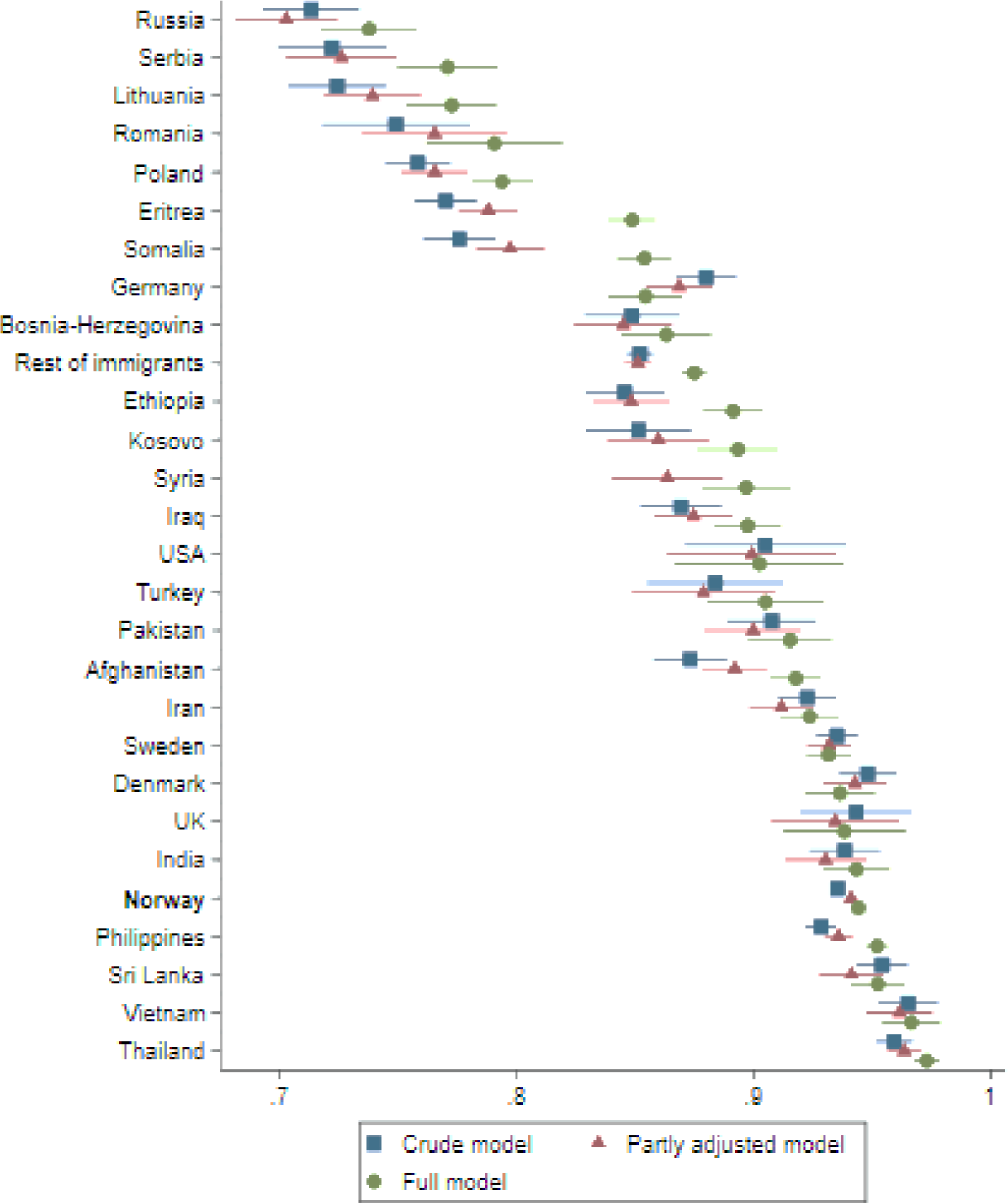
Predicted probability of COVID-19 vaccination by country of birth on 31 August 2021, with 95% confidence intervals, based on three logistic regression models: Crude, partly and fully adjusted. Reference category: Non-immigrants (Norway, bold). Partly adjusted model: adjusted for age, sex and municipality. Full model: adjusted for age, sex, municipality, COVID-19, risk group for severe COVID-19, occupation and health care service of employment. All regression results can be found in supplementary table 3.

Several of the immigrant groups have a higher vaccination rate compared to non-immigrants in the crude model (Vietnam, Thailand, Sri Lanka, Denmark, UK and India), but this finding does not hold for all of these immigrant groups in the full model. Only immigrants from Thailand (0.973) Vietnam (0.966), Sri Lanka (0.952) and The Philippines (0.952) are the immigrant groups that have a higher predicted probability than non-immigrants in the full model.

Several of the immigrant groups have a higher vaccination rate compared to non-immigrants in the crude model (Vietnam, Thailand, Sri Lanka, Denmark, UK and India), but this finding does not hold for all of these immigrant groups in the full model. Only immigrants from Thailand (0.973) Vietnam (0.966), Sri Lanka (0.952) and Philippines (0.952) are the immigrant groups that have a higher predicted probability than non-immigrants in the full model.

## Discussion

Overall, both immigrants and *Norwegian-born with immigrant parents* have lower vaccination rates than non-immigrants in the crude and adjusted models. Vaccination rates vary between immigrant groups based on country of birth. Some of the differences are likely to related to different occupational composition by country of birth.

The seven immigrant groups with the lowest vaccination rates displayed two distinctive patterns when all covariates were introduced. For five of the immigrant groups from Eastern Europe (Russia, Serbia, Lithuania, Romania and Poland), adjusting for compositional covariates did not explain much of the difference to rest of the population. Contrarily, for the two immigrant groups from East Africa with low vaccination rate (Eritrea and Somalia), we observe a considerable increase in the predicted probability of vaccination in the full model, suggesting that the lower vaccination rates in these groups can to a great degree be related to the occupational composition. The vaccination rate for health care assistants (89%) was lower than the other health care occupations (e.g. physicians (97%), nurses (93%)). In general, 25% of HCWs were health care assistants, while the share of HCWs from Eritrea and Somalia that were health care assistants were 50% and 43%. Other immigrant groups with a high percentage working as health care assistants, such as immigrants from Syria (65%) and Afghanistan (48%), showed a similar, although smaller, increase in the predicted probability when controlling for detailed occupation codes in the full model. For some of the Eastern European countries with lower vaccine rates occupational composition may be loosely related, but it does not appear to be as important as for Eritrea and Somalia. Therefore, there are likely to be other factors than the occupational composition related to the lower vaccine rates between immigrant groups.

A lower vaccination rate among health care assistants may be associated with both a lower education level and access. Supplementary figure 1 shows that health care assistants have had a lower vaccination rate than the other health care occupations during the entire period of vaccine roll-out, which is likely related to which occupations that have been prioritized first. Studies have also shown more vaccine hesitancy among groups with lower education level.^3^

Why certain immigrant groups have lower vaccine rates may be a result of multiple factors and barriers. Access to vaccines, or when they have been offered a vaccine, may play a role. The regression models have therefore aimed to capture some the most likely factors influencing HCWs’ likelihood to be offered a COVID-19 vaccine. The factors that have been emphasized in vaccine prioritization in Norway has been high age, prioritized HCWs, risk groups for severe illness from COVID-19, and municipalities with persistently high numbers of confirmed cases. These factors have been adjusted for - in addition to other factors that may influence either prioritization or the individual choice of accepting a vaccine, such as sex, COVID-19 infection, and type of health care service of employment. However, there may exist systematically differences in access between the immigrant groups that this study has not captured - e.g. language, logistic difficulties, lower digital competence, or lack of access to digital platforms.

Another barrier to vaccination may be vaccine hesitancy. Many of the immigrant groups with lower vaccination rates in this study have also shown a higher vaccine hesitancy rate in previous studies. Nilsen et al. found that persons born in Eastern Europe, Western Asia and Africa had the lowest rates of willingness to take the COVID-19 vaccine in Norway, with only 40% of the persons born in Eastern Europe being willing to take the COVID-19 vaccine.^2^ This finding on vaccine hesitancy largely corresponds to the vaccination rates disaggregated on country of birth in this study. On the opposite end of the scale, immigrants from countries in East Asia (Philippines, Sri Lanka, Thailand and Vietnam) and Nordic countries (Denmark and Sweden) displayed a high vaccine rate in our study, and had a correspondingly high willingness rate in Nilsen et al (71.74 and 71.17 respectively)^2^. This may suggest that vaccine hesitancy is one of the factors related to the differences in vaccine rate. Regardless of how important hesitancy is for differences in vaccination rate, the results demonstrate that the vaccination varies with country of birth. Consequently, for measures to improve vaccination uptake to be effective, they should focus on specific immigrant groups rather than all immigrants together.

There are a few limitations that may bring some uncertainty to the study. First, some immigrant HCWs may have been vaccinated in their country of birth but not registered as such in Norway. We do not know the full extent of this, and it may be a reason for a lower rate, especially for the Eastern European countries. Additionally, there are HCW without a permanent identity number, which cannot be included since they are not registered with country of birth. It means that the sample are not capturing all immigrant HCW in Norway, and it may create some systematic differences. However, it is not likely that the magnitude of these limitations may alter the main results.

Which of the factors or barriers that have contributed the most to a lower vaccine rate for some groups has not been examined in this study. If vaccine hesitancy is a barrier, we do not know how deeply ingrained it is, or how easily it can change over time with more information about the vaccines. Since the population sample of this study is HCWs, so we could assume that HCWs have better, or at least equal, willingness to take a COVID-19 vaccine than the overall population. To ensure the highest possible vaccination rates in all groups we need more knowledge about this topic - we therefore call for more research to understand barriers to vaccination, including vaccine hesitancy.

## Conclusion

Among HCWs, both immigrants (85%) and *Norwegian-born to immigrant parents* (88%) had lower vaccination rates than rest of the population (94%). There is a large variation between immigrant groups, with immigrants from Russia (71%), Serbia (72%), Lithuania (72%), Romania (75%), Poland (76%), Eritrea (77%), and Somalia (78%) having the lowest vaccination rate. Immigrants from Eritrea and Somalia more often worked as health care assistants, which in general had a lower vaccine rate than other health care occupations. When we therefore adjusted for demographics and detailed occupational codes the predicted probability of vaccination increased for Eritrea (0.849) and Somalia (0.854). The large variation between immigrant groups suggest that measures to improve vaccine uptake should focus on specific immigrant groups rather than all immigrants together. More research is required to investigate underlying reasons for differences in vaccination rates among HCWs with different country backgrounds.

## Supporting information

Supplementary figure 1: Percent vaccinated HCW over time by occupation in Norway 2021.

Supplementary table 1: Descriptive statistics of population sample by immigrant groups in Norway at 1 December 2020.

Supplementary table 2a and b: Standard Industrial Classification 2007 and Classification of Occupations 08.

Supplementary table 3: Logistic regression results of the effect of country of birth on August 2021.

## Data Availability

We used data from Beredt-C19, which was established by The Norwegian Institute of Public Health (NIPH) in cooperation with the Norwegian Directorate of Health and contains individual-level data for all Norwegian residents. This data is not public available.

## Funding

The authors received no external funding

## Conflict of interest

None declared

## Key points

- Immigrant HCW had a lower vaccination rate than HCWs without an immigrant background
- Vaccination rate varied country of birth, with immigrants from Eastern Europe and East Africa having the lowest vaccination rate
- The vaccination rate immigrants from Eritrea and Somalia increased when we adjusted for demographics and occupation, suggesting that occupational composition is related to the vaccination rate.
- The large variation between immigrant groups suggest that measures to improve vaccine uptake should focus on specific immigrant groups rather than all immigrants together.

## Notes

### Competing Interest Statement

The authors have declared no competing interest.

### Author Declarations

The establishment of an emergency preparedness register forms part of the legally mandated responsibilities of The Norwegian Institute of Public Health (NIPH) during epidemics. Institutional board review was conducted, and the Ethics Committee of South-East Norway confirmed (June 4th, 2020, #153204) that external ethical board review was not required. All necessary patient/participant consent has been obtained and the appropriate institutional forms have been archived.

