## Supplementary figure 1: Percent vaccinated HCW over time by occupation in Norway 2021. for "COVID-19 vaccination rates among health care workers by immigrant background. A nation-wide registry study from Norway"

Supplementary figure 1: Percent vaccinated HCW over time by occupation in Norway 2021. X-axis: calendar weeks in 2021.

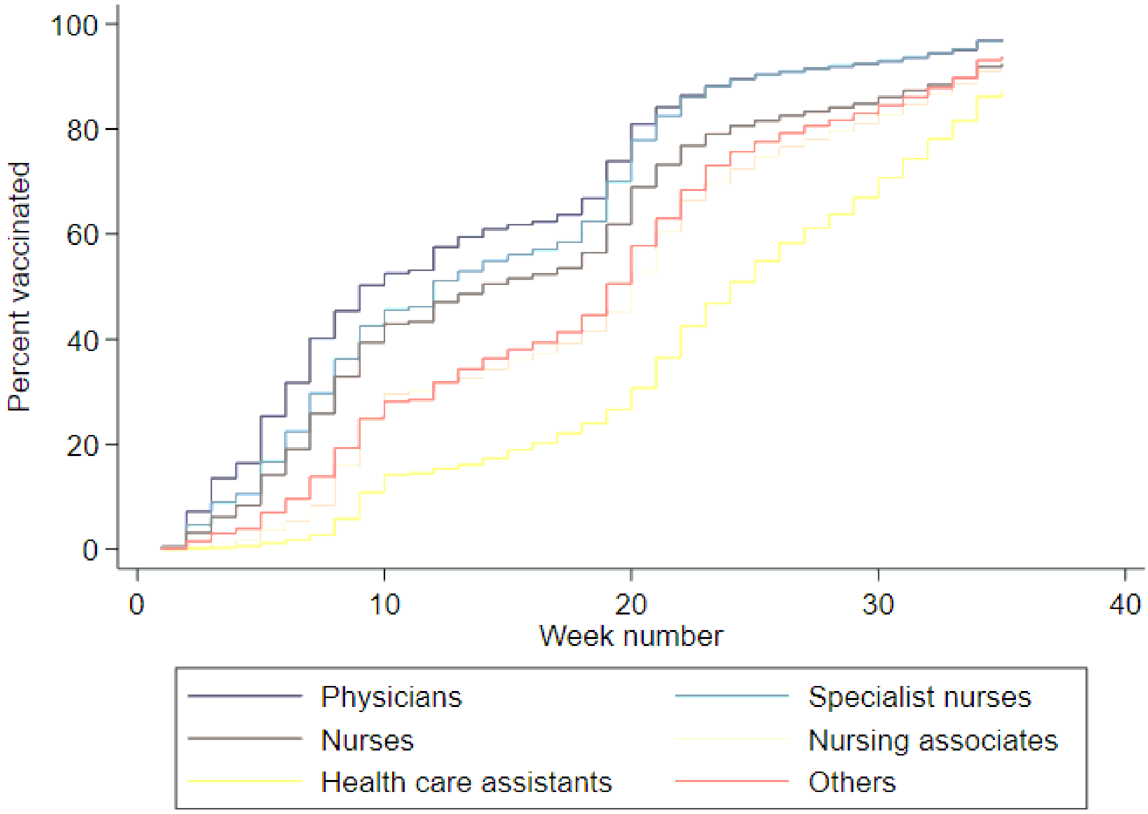
