## Supplementary table 1: Descriptive statistics of population sample by immigrant groups in Norway at 1 December 2020. for "COVID-19 vaccination rates among health care workers by immigrant background. A nation-wide registry study from Norway"

| Country of birth | N | Mean age (standard deviation) | Female, % | Living in Oslo, % |
|---|---|---|---|---|
| All | 356 053 | 41 (13) | 81 | 11 |
| Persons without immigrant background | 286 393 | 41 (13) | 82 | 09 |
| Immigrants | 62 339 | 41 (11) | 76 | 19 |
| Norwegian-born to immigrant parents | 5 477 | 29 (8) | 71 | 41 |
| Denmark | 1 281 | 47 (13) | 75 | 14 |
| Sweden | 2 972 | 44 (11) | 74 | 22 |
| Poland | 3 645 | 42 (10) | 89 | 11 |
| Romania | 730 | 41 (9) | 87 | 11 |
| Lithuania | 1 781 | 41 (10) | 92 | 07 |
| UK | 367 | 47 (12) | 57 | 15 |
| Russia | 1 863 | 44 (11) | 91 | 11 |
| Turkey | 473 | 41 (9) | 68 | 27 |
| Germany | 2 445 | 46 (11) | 70 | 10 |
| Bosnia-Herzegovina | 1 213 | 42 (10) | 75 | 21 |
| Serbia | 1 452 | 40 (9) | 64 | 23 |

| | | | | |
|---|---|---|---|---|
| Kosovo | 973 | 39 (11) | 75 | 15 |
| Eritrea | 3 956 | 37 (10) | 59 | 12 |
| Ethiopia | 1 791 | 39 (9) | 70 | 30 |
| Somalia | 2 878 | 33 (9) | 76 | 34 |
| Afghanistan | 1 745 | 32 (9) | 50 | 14 |
| Sri Lanka | 1 439 | 47 (10) | 68 | 38 |
| Philippines | 6 011 | 40 (9) | 85 | 23 |
| India | 972 | 44 (10) | 75 | 27 |
| Iraq | 1 464 | 37 (10) | 66 | 17 |
| Iran | 1 828 | 44 (11) | 64 | 27 |
| Pakistan | 901 | 40 (10) | 66 | 48 |
| Syria | 743 | 34 (9) | 41 | 09 |
| Thailand | 2 356 | 42 (10) | 96 | 06 |
| Vietnam | 792 | 44 (9) | 84 | 20 |
| USA | 283 | 45 (11) | 73 | 14 |
| Rest of countries | 16048 | 42 (11) | 77 | 17 |
