## Supplementary table 2a and b: Standard Industrial Classification 2007 and Classification of Occupations 08. for "COVID-19 vaccination rates among health care workers by immigrant background. A nation-wide registry study from Norway"

Supplementary table 2a: Standard Industrial Classification 2007 (SIC 2007)

| **Code** | **Industry** |
|---|---|
| 86.1 | Hospital activities |
| 86.2 | Medical and dental practice activities |
| 86.901 | Home nursing care |
| 86.902 | Physiotherapy services |
| 86.903 | Maternal and child health care and school health services |
| 86.906 | Medical laboratories |
| 86.907 | Ambulance services |
| 87.1 | Residential nursing activities |
| 87.2 | Residential care activities for mental retardation, mental health and substance abuse |
| 87.3 | Residential care activities for the elderly and disabled |
| 88.1 | Social work activities without |

| | |
|---|---|
| | accommodation for the elderly and disabled |

Supplementary table 2b: Classification of Occupations 08 (STYRK 08) [1]

| Codes | Occupation |
|---|---|
| 221 | Medical doctors |
| 222 | Nursing and midwifery professionals |
| 226 | Other health professionals |
| 2634 | Psychologists |
| 3211 | Medical imaging and therapeutic equipment technicians |
| 3212 | Medical and pathology laboratory technicians |
| 3251 | Dental assistants and therapists |

[1] If a person was registered with several employment contracts, we have selected the occupational contract based on the following order: Physicians, specialist nurses, midwifes, nurses, social educators, dentists, physiotherapists, occupational therapists, psychologists, radiographers (and associated staff, bioengineers, medical assistants, ambulance personnel, nursing associates, health care assistants, cleaners and other health care workers

| | |
|---|---|
| 3254 | Dispensing opticians |
| 3256 | Medical assistants |
| 3258 | Ambulance workers |
| 5321 | Health care assistants |
| 5322 | Home-based personal care workers |
| 5329 | Personal care workers in health services not elsewhere classified |
| 9112 | Cleaners and helpers in offices, hotels and other kinds of activity units |
