## Supplementary table 3: Logistic regression results of the effect of country of birth on August 2021. for "COVID-19 vaccination rates among health care workers by immigrant background. A nation-wide registry study from Norway"

Supplementary table 3: Logistic regression results of the effect of country of birth on August 2021. Odds ratio, t-statistics in parenthesis. Three logistic regression models are presented: Crude, partly and fully adjusted. Reference category: Non-immigrants (Norway, bold). Partly adjusted model: adjusted for age, sex and municipality. Full model: adjusted for age, sex, municipality, COVID-19, risk group for severe COVID-19, occupation and health care service of employment.

| Variables | Crude model | Partly adjusted model | Full model |
|---|---|---|---|
| | Odds ratio (t-statistics) | Odds ratio (t-statistics) | Odds ratio (t-statistics) |
| Country of birth | | | |
| *Norway* | 1 | 1 | 1 |
| | (.) | (.) | (.) |
| *Denmark* | 1.262 | 1.030 | 0.870 |
| | (1.85) | (0.23) | (-1.08) |
| *Sweden* | 0.996 | 0.856* | 0.806** |
| | (-0.05) | (-2.07) | (-2.84) |
| *Poland* | 0.217*** | 0.205*** | 0.228*** |
| | (-38.79) | (-38.90) | (-35.20) |
| *Romania* | 0.206*** | 0.205*** | 0.224*** |
| | (-18.46) | (-18.10) | (-16.58) |

| | | | |
|---|---|---|---|
| *Lithuania* | 0.182*** | 0.178*** | 0.201*** |
| | (-31.99) | (-31.38) | (-28.30) |
| *UK* | 1.149 | 0.893 | 0.900 |
| | (0.62) | (-0.50) | (-0.46) |
| *Russia* | 0.172*** | 0.148*** | 0.167*** |
| | (-34.09) | (-35.62) | (-32.52) |
| *Turkey* | 0.525*** | 0.454*** | 0.564*** |
| | (-4.48) | (-5.44) | (-3.88) |
| *Germany* | 0.506*** | 0.414*** | 0.347*** |
| | (-10.90) | (-13.79) | (-16.17) |
| *Bosnia-Herzegovina* | 0.387*** | 0.342*** | 0.373*** |
| | (-11.83) | (-13.15) | (-11.85) |
| *Serbia* | 0.179*** | 0.166*** | 0.199*** |
| | (-29.10) | (-29.49) | (-25.54) |
| *Kosovo* | 0.396*** | 0.384*** | 0.496*** |
| | (-10.27) | (-10.46) | (-7.40) |
| *Eritrea* | 0.231*** | 0.233*** | 0.332*** |

| | | | |
|---|---|---|---|
| | (-38.00) | (-36.97) | (-27.11) |
| *Ethiopia* | 0.378*** | 0.351*** | 0.485*** |
| | (-14.78) | (-15.82) | (-10.70) |
| *Somalia* | 0.239*** | 0.247*** | 0.346*** |
| | (-31.62) | (-30.37) | (-22.19) |
| *Afghanistan* | 0.475*** | 0.518*** | 0.660*** |
| | (-10.32) | (-8.99) | (-5.60) |
| *Sri Lanka* | 1.437** | 1.004 | 1.184 |
| | (2.87) | (0.03) | (1.32) |
| *Philippines* | 0.896* | 0.914 | 1.175** |
| | (-2.17) | (-1.75) | (3.07) |
| *India* | 1.052 | 0.837 | 0.985 |
| | (0.38) | (-1.32) | (-0.11) |
| *Iraq* | 0.460*** | 0.437*** | 0.518*** |
| | (-10.02) | (-10.59) | (-8.31) |
| *Iran* | 0.823* | 0.647*** | 0.716*** |
| | (-2.23) | (-4.95) | (-3.72) |

| | | | |
|---|---|---|---|
| *Pakistan* | 0.679*** | 0.562*** | 0.640*** |
| | (-3.37) | (-4.97) | (-3.75) |
| *Syria* | 0.363*** | 0.397*** | 0.515*** |
| | (-10.10) | (-9.02) | (-6.40) |
| *Thailand* | 1.626*** | 1.653*** | 2.129*** |
| | (4.65) | (4.78) | (7.11) |
| *Vietnam* | 1.913*** | 1.562* | 1.701** |
| | (3.37) | (2.32) | (2.73) |
| *USA* | 0.658* | 0.559** | 0.547** |
| | (-2.07) | (-2.90) | (-2.97) |
| *Remaining Immigrants* | 0.398*** | 0.358*** | 0.414*** |
| | (-39.29) | (-42.44) | (-35.21) |
| Age | | | |
| *<29 years old* | | 1 | 1 |
| | | (.) | (.) |
| *30-39 years old* | | 1.022 | 0.831*** |
| | | (1.34) | (-10.54) |

| | | |
|---|---|---|
| *40-49 years old* | 1.793$^{***}$ | 1.454$^{***}$ |
| | (30.57) | (18.37) |
| *50-59 years old* | 2.135$^{***}$ | 1.799$^{***}$ |
| | (36.59) | (26.88) |
| *>60 years old* | 2.434$^{***}$ | 2.030$^{***}$ |
| | (28.36) | (21.98) |
| Female | 0.806$^{***}$ | 0.812$^{***}$ |
| | (-12.41) | (-11.60) |
| COVID-19 | | 0.486$^{***}$ |
| | | (-26.45) |
| Occupation | | |
| *Physicians* | | 1 |
| | | (.) |
| *Specialist nurses* | | 0.760$^{***}$ |
| | | (-5.01) |
| *Midwifes* | | 0.417$^{***}$ |
| | | (-8.91) |

staff)

| | |
|---|---|
| *Nurses* | 0.422$^{***}$ |
| | (-19.12) |
| *Social educators* | 0.446$^{***}$ |
| | (-15.16) |
| *Dentists* | 0.557$^{***}$ |
| | (-5.88) |
| *Physiotherapists* | 0.792$^{**}$ |
| | (-2.90) |
| *Occupational therapists* | 0.761$^{*}$ |
| | (-2.53) |
| *Psychologists* | 0.791$^{**}$ |
| | (-2.69) |
| *Radiographers (and associated staff)* | 0.964 |
| | (-0.32) |

| | |
|---|---|
| *Bioengineers* | 0.791** |
| | (-2.91) |
| *Medical assistants* | 0.393*** |
| | (-14.98) |
| *Ambulance personnel* | 0.924 |
| | (-0.90) |
| *Nursing associates* | 0.377*** |
| | (-21.55) |
| *Health care assistants* | 0.332*** |
| | (-24.50) |
| *Cleaners* | 0.286*** |
| | (-21.05) |
| *Other HCW* | 0.437*** |
| | (-7.90) |
| Institution of | |

| | |
|---|---|
| Employment | |
| *Hospital* | 1 |
| | (.) |
| *Other specialist health service* | 0.670*** |
| | (-12.30) |
| *Primary health service (excl. nursing homes)* | 0.813*** |
| | (-5.64) |
| *Home care service* | 0.568*** |
| | (-21.29) |
| *Nursing home* | 0.655*** |
| | (-16.32) |
| *Other health service* | 0.568*** |
| | (-21.97) |
| Risk group of severe COVID- | 1.285*** |

| | | | |
|---|---|---|---|
| | | | (11.57) |
| *N* | 355893 | 355893 | 355893 |

Odds ratio; *t* statistics in parentheses: $^{*}\ p < 0.05$, $^{**}\ p < 0.01$, $^{***}\ p < 0.001$. Likelihood ratio tests ran on models:  the partly adjusted model is a significant improvement of the crude model (p<0.00), and the full model is a significant improvement of the partly adjusted model (p<0.00).
